# Age- and airway disease related gene expression patterns of key SARS-CoV-2 entry factors in human nasal epithelia

**DOI:** 10.1101/2021.05.23.21257673

**Authors:** Mihkel Plaas, Kadri Seppa, Nayana Gaur, Priit Kasenõmm, Mario Plaas

**Affiliations:** Tartu University Hospital, Ear Clinic (Kuperjanovi 1, 51003 Tartu, Estonia); University of Tartu, Institute of Clinical Medicine. (L. Puusepa 8, 50406 Tartu, Estonia); University of Tartu, Institute of Biomedicine and Translational Medicine, Department of Physiology. (Ravila 19, 50411 Tartu, Estonia); Friedrich Schiller University Jena, Hans Berger Department of Neurology. (Am Klinikum 1, 07747 Jena, Germany); University of Tartu, Institute of Biomedicine and Translational Medicine, Laboratory Animal Centre. (Ravila 14B, 50411 Tartu, Estonia)

**Keywords:** SARS-CoV-2, ACE2, AGTR2, NRP1, TMPRSS2

## Abstract

The global COVID-19 pandemic caused by SARS-CoV-2 predominantly affects the elderly. Differential expression of SARS-CoV-2 entry genes may underlie the variable susceptibility in different patient groups. Here, we examined the gene expression of key SARS-CoV-2 entry factors in mucosal biopsies to delineate the roles of age and existing chronic airway disease. A significant inverse correlation between *ACE2* and age and a downregulation of NRP1 in patients with airway disease were noted. These results indicate that the interplay between various factors may influence susceptibility and the disease course.

## 1. Introduction

A novel coronavirus (SARS-CoV-2) was first identified in Wuhan, China in November 2019 and has since spread worldwide causing the current COVID-19 pandemic [1, 2]. The disease mostly affects the elderly [3] and as of January 2021, it had caused more than 1.9 million deaths globally [2]. SARS-CoV-2 enters the upper airways via the nasal epithelium and, like other coronaviruses, uses the *ACE2* receptor for cellular entry. Other co-receptors, like *TMPRSS2* also reportedly facilitate viral entry [4, 5]. While both *ACE2* and *TMPRSS2* are expressed in a broad range of tissues, co-expression only occurs in certain cellular populations, indicating that other receptors may mediate viral susceptibility [4, 6]. This is reinforced by clinical observations of a discrepancy between affected organs and *ACE2* expression and the fact that not all cells with *ACE2/TMPRSS2* co-expression are susceptible to the virus [7].

*AGTR2* (angiotensin II receptor type 2) may be a potential co-entry factor as it was shown to have a higher binding affinity to SARS-CoV-2 spike (S) protein than *ACE2* in a 3D structural simulation [8], although this is yet to be confirmed with in/ex-vivo experimental data. Neuropilin-1 (NRP1) has also been shown to potentiate SARS-CoV-2 entry and host-cell susceptibility [9, 10]. Following cleavage of the Spike (S) protein, a polybasic Arg-Arg-Ala-Arc C terminal sequence is generated which can then bind to the cell surface via the *NRP1* receptor [9]. High *NRP1* expression has been reported in pulmonary and olfactory cells, with the highest expression in endothelial cells. In addition, *NRP1* may be an attractive entry factor for viruses owing to its high expression on epithelia in contact with external environment [10].

Although additional factors, including age and the presence of chronic airway disease (including for instance asthma and chronic rhinosinusitis (CRS)) might influence susceptibility to COVID-19, their contribution is yet to be rigorously studied and results have been inconsistent. For instance, while Bunyavanich et al [11], reported that *ACE2* expression in nasal epithelia increases with age, similar studies observed no association between age and *ACE2* expression [12]. Lower *ACE2* expression levels have also been noted in patients with asthma [13], while lower levels of *TMPRSS2* were noted in patients with chronic rhinosinusitis (CRS) [14]. These observations might suggest altered susceptibility for COVID-19 in these patient groups.

At present, there are relatively few studies that have comprehensively examined the roles of age and airway disease on the expression of key SARS-CoV-2 entry factors. Therefore, the objectives of the present study were to 1) investigate the expression of key entry factors *ACE2, TMPRSS2, NRP1* and *AGTR2* and 2) delineate the influence of age and chronic airway disease within a single clinically characterized cohort.

## 2. Methods

This study enrolled 49 patients across all age groups who were consecutively recruited between September and November 2020 from the Department of Otolaryngology at the Tartu University Hospital. The study was approved by the Research Ethics Committee of the University of Tartu (approval nr 322/T-3) and written informed consent was obtained from all patients and/or their representatives. Biopsies were collected from the inferior turbinate during the course of elective nasal and/or nasopharyngeal surgeries and stored at -80 °C until further processing. Subjects with autoimmune, metabolic or oncologic disease were excluded. Within children, the most common reason for surgery was adenoidectomy (94%), while within adults it was septoplasty (54%) or endoscopic sinus surgery (40%). Eight adult patients (16%) had hypertension and were receiving medication. Biopsies were homogenized using the Precellys CK14 Lysing Kit and the Precellys homogenizer (both Bertin Instruments, USA). RNA was isolated using the Direct-zol RNA MiniPrep Kit in accordance with manufacturer instructions (Zymo Research). SuperScript III Reverse Transcriptase (Invitrogen) and random hexamers were used to synthesize cDNA according to the manufacturer’s protocol. All RT-qPCR reactions were performed using Taqman Gene Expression MasterMix (Thermo Fisher Scientific) and TaqMan Gene Expression Assays. The 2-ΔCt method was used for relative quantification of gene expression; expression was calculated relative to the internal control *HPRT1*. TaqMan probe details are provided in Supplementary Table 1.

Statistical analyses were performed using the SPSS (Version 26; IBM Corp.) and GraphPad Prism (version 9.0.0 for Windows) software packages. The Shapiro-Wilk test was used to check for data normality. Correlations between continuous variables were assessed using the Spearman’s test. Between-group comparisons for chronic airway disease (defined as having asthma and/or CRS in history) vs. no airway disease group were performed using either the independent samples t-test or the Mann-Whitney U test. A one-way ANOVA (with post-hoc Tukey test) was performed to assess the effect of age on target gene expression. Assumptions for sphericity and homogeneity of variances and co-variances were met unless stated otherwise. All outliers were retained for analyses. Summary data are reported as the mean with either 95% confidence intervals (CI) or the standard deviation (SD). All analyses were performed on fold changes calculated using the 2-ΔCt method. Two-tailed statistical significance was set at p < 0.05.

## 3. Results

While 49 samples were initially collected, the final cohort included 46 individuals; three were excluded owing to insufficient RNA amount. The cohort age ranged from 2-66 years. Further analyses were performed by stratifying the cohort based on either 1) age or 2) history of chronic airway disease. For the age sub-division, the cohort was divided into 3 age groups (Table 1). Normality testing for all continuous variables showed that age and *AGTR2* expression levels were non-normally distributed (p < 0.05). Sex distribution was equally split across all 3 age groups.

**Table 1:** Gene expression comparison between age groups

|  | **Children (0-17)** | **Young adults**<br>**(18-39)** | **Older adults**<br>**(40-66)** | **p-value** |
| --- | --- | --- | --- | --- |
| **N** | 18 | 15 | 13 |  |
| **Age range (mean, SD)** | 2-13 (4.9 ± 2.9) | 21-39 (30.2 ± 5.5) | 40-66 (56.6 ± 8.7) |  |
| **Sex distribution (% of males)** | 8 (44%) | 8 (53%) | 7 (53%) |  |
| ***ACE2*** | 0.56 (± 0.18) | 0.36 (± 0.22) | 0.30 (± 0.19) | 0.002 |
| ***TMPRSS2*** | 3.12 (± 1.14) | 4.01 (± 1.28) | 3.19 (± 1.41) | 0.112 |
| ***AGTR2*** | 0.14 (± 0.36) | 0.11 (± 0.21) | 0.031 (± 0.025) | 0.590 |
| ***NRP1*** | 4.91 (± 1.76) | 4.47 (± 1.97) | 3.86 (± 1.19) | 0.253 |

Gene expressions are given as mean (SD) fold-change values.

For the chronic airway disease sub-division, the cohort was divided into 2 sub-groups (Table 2); no significant differences in sex distribution or age were noted (p = 0.073; t = 1.273).

**Table 2:** Gene expression comparison between airway disease (asthma, CRS) and no airway disease groups

|  | No airway disease | Airway disease | p-value |
| --- | --- | --- | --- |
| **N** | 33 | 13 |  |
| **Age range (mean, SD)** | 2-66 (25.3 ± 20.2) | 3-65 (33.8 ± 25.6) |  |
| **Sex distribution (% of males)** | 17 (51%) | 6 (46%) |  |
| ***ACE2*** | 0.46 (± 0.23) | 0.32 (± 0.19) | 0.074 |
| ***TMPRSS2*** | 3.43 (± 1.28) | 3.42 (± 1.42) | 0.969 |
| ***AGTR2*** | 0.049 (± 0.10) | 0.23 (± 0.43) | 0.321 |
| ***NRP1*** | 4.78 (± 1.81) | 3.67 (± 1.14) | 0.047 |

Gene expressions are given as mean (SD) fold change values.

### 3.1 Age Group Stratification

While no significant differences were noted for *AGTR2, TMPRSS2* or *NRP1* expression, *ACE2* expression significantly differed between age sub-groups (p = 0.002; F (2, 43) = 7.514), as seen in Table 1. Post-hoc analysis showed a significant decrease in *ACE2* expression levels in both younger (0.201, 95% CI [0.029, 0.372], p = 0.01) and older adults (0.265, 95% CI [0.086, 0.444], p = 0.002) compared to those in children, as seen in Figure 1. Correlation analysis for the entire cohort revealed a significant inverse correlation between age and *ACE2* expression (Spearman’s rho r = -0.561, p < 0.001).

**Figure 1.**
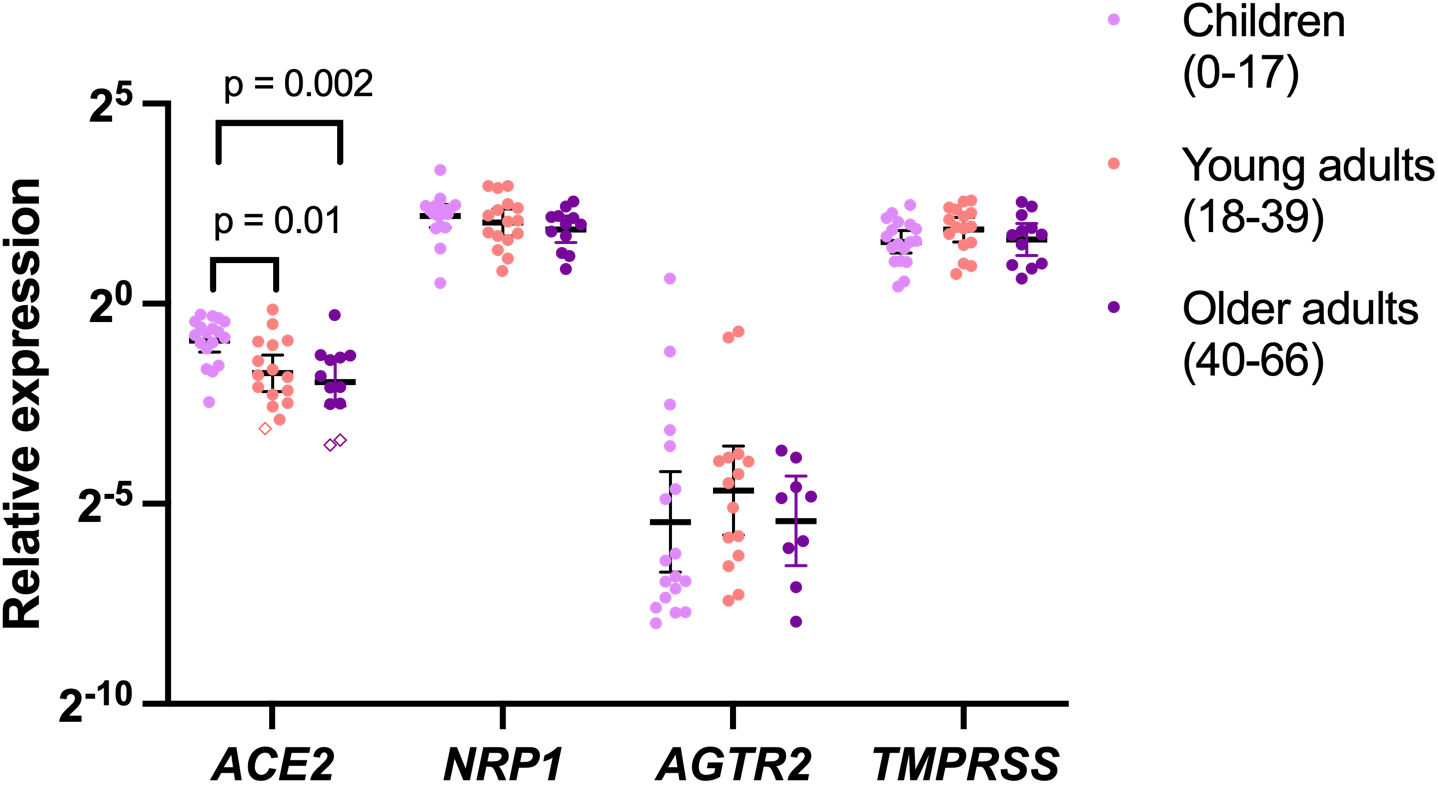
Between age-group comparisons of key SARS-CoV-2 entry factors. Differences in the relative expression of *ACE2, TMPRSS2, AGTR2* and *NRP1* between age subgroups is displayed. Between group differences were calculated using a one-way ANOVA with post-hoc Tukey test. Data is presented as individual scatterplots with the geometric mean and 95% confidence intervals. P values are reported for statistically significant results. Y axis is displayed in the log2 scale. Symbols presented as ♦ indicate individuals who were subsequently diagnosed with COVID-19.

### 3.2 Disease Stratification

No statistically significant differences were noted for *ACE2, TMPRSS2* or *AGTR2* expression between airway disease and no-airway disease groups. Interestingly, *NRP1* expression was significantly lower in the airway disease group (3.67, SD = 1.14) relative to the no-disease group (4.78, SD = 1.81) (p < 0.05), as seen in Figure 2. However, after controlling for age as a co-variate, this result did not retain statistical significance (p = 0.08).

**Figure 2.**
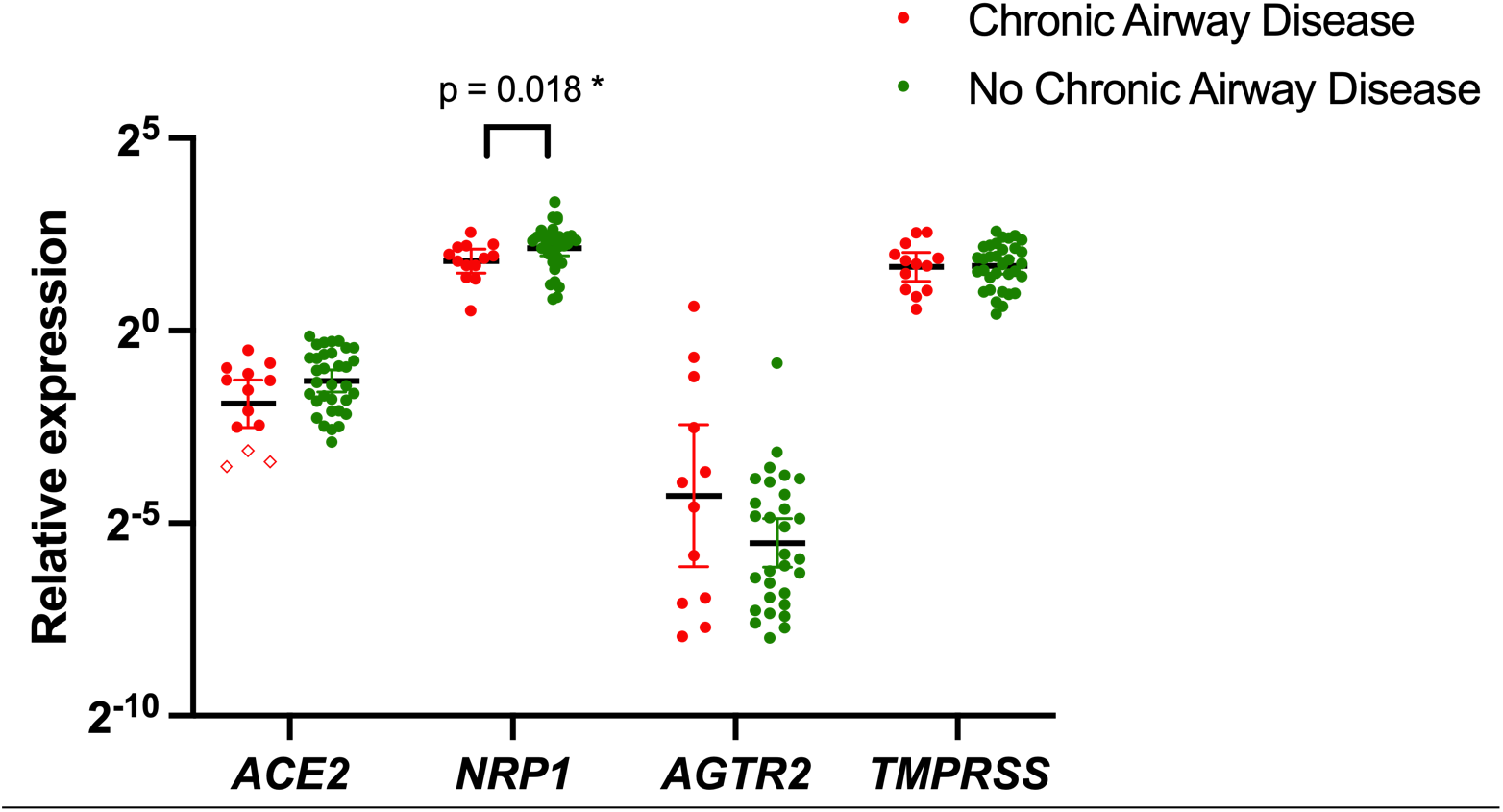
Between airway disease-group comparisons of key SARS-CoV-2 entry factors. Differences in the relative expression of *ACE2, TMPRSS2, AGTR2* and *NRP1* between airway disease and no disease groups are displayed. Between-group comparisons were performed using either the independent samples t-test or the Mann-Whitney U test. Data is presented as individual scatterplots with the geometric mean and 95% confidence intervals. P values are reported for statistically significant results. Y axis is displayed in the log2 scale. Symbols presented as ♦ indicate individuals who were subsequently diagnosed with COVID-19. *\* this value did not retain statistical significance after controlling for age as a covariate*.

## 4. Discussion

Despite the changing demographics of COVID-19 and increased infection rates in the young population, mortality rates are primarily driven by the elderly. For example, in the United States, while people over 50 years account for ∼35% of the total case number, they represent 95% of total fatalities. In contrast, children accounted for only 0.2% of total deaths as of January 2021 [3]. Age-related differences in the expression of SARS-CoV-2 entry factors, in particular *ACE2*, may underlie these distinctive susceptibility profiles. In contrast to Bunyavanich *et al*. [11], we found that *ACE2* levels are inversely correlated with age. This observation is also concordant with those reported by other, more extensive studies. For instance, Chen *et al*. noted a cohort-wide age-associated decrease in *ACE2* expression following analysis of the Genotype-Tissue Expression (GTEx) project dataset [15]. These divergent results may be explained by the somewhat paradoxical protective role of *ACE2* in COVID-19 pathology. Previous studies demonstrate that children have a reduced risk of infection for SARS-CoV-2 despite having higher *ACE2* levels. *ACE2* levels are lowest in the elderly i.e. the population demographic most at risk for COVID-19. While increased *ACE2* expression may initially facilitate infection, it conversely also confers protection against the subsequent inflammatory cascade, indicating the duality of its role in COVID-19 pathology. This protective effect is primarily driven through two different *ACE2* functions. First, *ACE2* counteracts the vasoconstrictive and inflammatory actions of angiotensin 2 and maintains a functioning *ACE2* - Angiotensin – (1-7) – MAS system. Indeed, the loss of physiological *ACE2* function skews affected cells towards a pro-apoptotic state and reduces cell survival-associated signalling. Secondly, circulating soluble *ACE2* enzyme can bind to SARS-CoV-2, therefore competitively reducing the amount of virus that can potentially bind to the membrane-localized *ACE2* receptor. The increased *ACE2* expression noted in children in the present study may therefore also explain why they are typically asymptomatic and/or have a much milder disease course [16-19].

It is worth noting that conflicting results between the present and previous studies may also be due to methodological differences, in particular sample collection and processing e.g. cytology brush samples vs mucosal biopsies.

Of note, three subjects in our cohort eventually contracted COVID-19. Curiously, these three individuals also had the lowest values observed for *ACE2* expression within this cohort, further suggesting that higher *ACE2* expression may confer protection against COVID-19 (individual values indicated in Figure 1 and 2). No other relevant expression trends for the other gene targets were noted. Naturally however, the limited sample size and statistical power preclude any quantitative analyses and we present this as a retrospective observation.

No age-associated differences in expression were noted for the other receptors investigated in this study, which is in keeping with previous reports. However, we found that the presence of chronic airway disease is associated with lower *NRP1* expression. Disease-associated downregulation has also been reported for *ACE2* and *TMPRSS2* in the presence of asthma and CRS [13, 14]. While no significant disease-associated changes in *TMPRSS2* expression were noted in the present study, this may be because of the aforementioned sampling and methodological differences. Although the downregulation of SARS-CoV-2 entry genes might suggest that these patients are less susceptible to disease, our clinical statistics show that prevalence rates within hospitalized COVID-19 patients who had either asthma, CRS or both, were similar to those within the general population.

With regard to other comorbidities, we observed that mean *ACE2* expression was lower in individuals with hypertension; however, this effect did not remain significant after the inclusion of age as a co-variate *(data not shown)*. Although no other correlations were noted between gene expression and clinical indices, this may also be a result of limited sample size.

## 5. Conclusion

To summarize, while there is already a sizeable body of literature on SARS-CoV-2, its cellular entry mechanisms remain to be fully clarified; existing reports do not fully explain the clinical pattern of COVID-19. The present study uses a clinically characterized cohort to demonstrate how age, via its effect on putative entry receptors, can influence the disease course. Our data also reiterates the central role of *ACE2* in mediating COVID-19 pathology and highlights its paradoxical role, wherein lower levels may potentially increase susceptibility.

An additional strength of the study is its use of full mucosal biopsies as these are more physiologically representative of the upper airway and its complex cellular milieu. Limitations include the restricted sample size, single centre design and the fact that analyses were restricted to transcriptomic expression only. We recommend that future studies confirm if the results observed here extend to the protein level, assess expression in confirmed COVID-19 patients, undertake a comprehensive evaluation of associated risk factors and further clarify the role of *NRP1* in COVID-19 pathophysiology.

## Data Availability

Supplementary data available per request

## Funding and acknowledgments

This research was supported by University of Tartu Laboratory Animal Centre and by grant PSG471 (Mario Plaas) from Estonian Research Council. NG is supported by a doctoral scholarship (Landesgraduiertenstipendien) from the Graduate Academy of Friedrich Schiller University, Jena, Germany and the state of Thuringia.

## Author contributions

Mar.P. conceived and directed the study. Mih.P., P.K and Mar. P. designed the experiments. K.S. performed gene expression analysis. Mih.P. and N.G. participated in analysis and interpretation of the data and writing of the manuscript. Mih.P. and N.G. compiled the manuscript. All the authors have read and approved the final version of the manuscript.

### List of Abbreviations

ACE2: Angiotensin-converting enzyme 2
TMPRSS2: Transmembrane protease, serine 2
AGTR2: Angiotensin II Receptor Type 2
NRP1: Neuropilin 1
CRS: Chronic rhinosinusitis

